# HAND HYGIENE, COMPLIANCE AND ALCOHOL-BASED PRODUCTS: WHAT WAS THE LEGACY OF THE COVID-19 PANDEMIC?

**DOI:** 10.1101/2025.09.09.25335069

**Authors:** Amanda Carina Coelho de Morais, Sílvia Maria dos Santos Saalfeld, César Helbel, Matheus Cordeiro Marchiotti, Hilton Vizi Martinez, Josy Anne Silva, Fernanda Cristina Coelho Musse, Maria Cristina Bronharo Tognim

## Abstract

In the era of multidrug-resistant organisms and following the coronavirus disease 2019 (COVID-19) pandemic, the prevention of healthcare-associated infections (HAIs) has become one of the most critical global health concerns. The World Health Organization (WHO) developed the “My Five Moments for Hand Hygiene,” encouraging healthcare professionals to improve adherence to hand hygiene (HH), thereby increasing compliance rates and emphasizing alcohol-based hand rubs as the preferred method. We investigated HH compliance and the use of alcohol-based formulations among healthcare professionals working in adult, pediatric, and neonatal intensive care units (ICUs), comparing the pre and post-pandemic periods. In the post-pandemic period, we analyzed HH compliance across ICUs as well as HH practices in each of the WHO’s “Five Moments.” A total of 2,789 HH opportunities were recorded (1,048 before the pandemic and 1,741 after). Overall compliance rates increased from 61% (640/1,048) before the pandemic to 66% (1,157/1,741) after, a statistically significant difference (p = 0.004). HH compliance is most critical in the moments before patient contact or aseptic procedures, especially in the Adult and Pediatric ICUs. The Neonatal ICU showed superior performance overall, including significantly higher performance during the two most critical periods. The results reinforce that the COVID-19 pandemic led to an increase in healthcare professionals’ HH compliance, although disparities persist between professional categories and hospital sectors. Furthermore, there was a drastic change in the pattern of selective hand sanitizer use, increasing the likelihood of alcohol use more than fourfold compared to the previous period (OR 4.30; 95% CI 3.32–5.58; p < 0.001).

## INTRODUTION

In the era of multidrug-resistant organisms, and especially after the coronavirus disease 2019 (COVID-19) pandemic, the prevention of healthcare-associated infections (HAIs) has become one of the most critical global health issues [1, 2, 3, 4]. Despite imprecise estimates regarding the full extent of the problem, current evidence indicates that millions of people worldwide suffer disabling harm or die due to failures in healthcare delivery, highlighting patient safety as a global public health concern [5, 6].

In Europe, this corresponds to 3.2 million individuals affected by HAIs in acute care hospitals annually, contributing to approximately 37,000 deaths [7]. In the United States, nearly 1 in every 25 hospitalized patients in intensive care units acquires an HAI [8]. Within this context, hand hygiene (HH) is recognized as the single most important procedure for preventing hospital infections [5], since many types of infections are caused by microorganisms transmitted via the contaminated hands of healthcare workers, particularly when HH is omitted, neglected, or inadequately performed [6, 9].

In recent years, healthcare workers’ HH compliance practices has been promoted by the World Health Organization (WHO), which launched in 2005 the “Multimodal Hand Hygiene Improvement Strategy” [10], including recommendations and interventions through multiple tools: system change, training, observation and feedback, workplace reminders, and institutional safety climate reinforcement. In 2008, the WHO strengthened this initiative with the global program “Clean Care is Safer Care,” aimed at improving HH compliance among healthcare professionals [11, 12].

Alongside these interventions, the WHO developed the “My Five Moments for Hand Hygiene” [6], encouraging healthcare professionals to systematically perform HH at five critical moments: (1) before touching a patient; (2) before clean/aseptic procedures; (3) after exposure risk to body fluids; (4) after touching a patient; and (5) after touching patient surroundings. This strategy is based on the conceptual model of cross-transmission of microorganisms and was designed to be applied in teaching, auditing, and reporting HH behavior [13].

During the COVID-19 pandemic, the importance of HH among healthcare workers was highlighted, and stricter infection prevention and control measures were implemented worldwide. For instance, the use of alcohol-based hand rubs as the preferred method was strongly recommended [13]. Consequently, it is expected that the pandemic experience has contributed to increased HH compliance and to the use of alcohol-based formulations among healthcare workers.

We investigated HH compliance among healthcare professionals working in adult, pediatric, and neonatal intensive care units (ICUs) in a teaching hospital, comparing pre and post-pandemic periods. We assessed the preference for different HH products, including non-medicated liquid soap, chlorhexidine, and alcohol-based preparations, before and after the pandemic. Finally, we analyzed current data regarding differences in HH compliance across the ICUs, as well as adherence to each of the WHO’s “Five Moments for Hand Hygiene.”

## METHODS

We performed a database analysis of the Hospital Infection Control Service of our Teaching Hospital, prior to the COVID-19 pandemic, between 09/01/2015 and 08/01/2018, assessing HH compliance among healthcare professionals working in Adult, Pediatric, and Neonatal Intensive Care Units (ICUs). Adherence was evaluated before and after contact with patients or equipment and considered adequate when hand rubbing was performed according to the technique recommended by the Brazilian Ministry of Health. The type of product used and the category of healthcare professional were also assessed. The second data survey was after the start of the 2019 coronavirus pandemic, between 10/29/2021 and 12/27/2024, evaluating HH compliance among healthcare professionals in Adult, Pediatric, and Neonatal ICUs for each of the WHO’s “Five Moments for Hand Hygiene.” The type of product used and the category of healthcare professional were also recorded. We used “IBM SPSS Statistics” for statistical analysis. To compare HH compliance rates before and after the pandemic (overall and by professional category), we applied the z-test for two independent proportions. For the analysis of the type of preparation used (non-medicated liquid soap or chlorhexidine vs. alcohol-based preparations), the chi-square test of independence was applied. We calculated the odds ratio (OR) with 95% confidence intervals (95% CI) as a measure of effect to quantify the association between the observation period (before vs. after the pandemic) and the type of preparation used. A significance level of p < 0.05 was adopted for all analyses. For the analysis of the WHO’s “Five Moments,” we applied Pearson’s chi-square test of independence, reporting p-values and Cramer’s V effect size. When the test was significant (Moments 1 and 2), we performed pairwise comparisons between ICUs (Adult vs. Pediatric, Adult vs. Neonatal, Pediatric vs. Neonatal) using two-tailed Fisher’s exact test with Holm adjustment. The study is part of the project “Strategies for reducing hospital costs and optimising the use of antimicrobials: rapid detection of resistance genes, synergism, and pharmacokinetic/pharmacodynamic analysis, phase 2” and was approved by the Permanent Research Ethics Committee Involving Human Beings of the State University of Maringá (CAAE 63610816.0.0000.0104).

## RESULTS

A total of 2,789 hand hygiene (HH) opportunities were recorded, with 1,048 before the COVID-19 pandemic and 1,741 after. The overall HH compliance rate before the pandemic was 61% (640/1,048) and 66% (1,157/1,741) after. Comparative analysis of HH compliance before and after the pandemic showed a significant increase in the overall rate (p = 0.004). When analyzing intensive care units (ICUs) separately, a substantial increase in adherence was observed in the Adult ICU, from 49.54% (164/331) to 62.88% (581/924) (p < 0.001), and in the Neonatal ICU, from 65.29% (237/363) to 79.49% (314/395) (p < 0.001). In contrast, the Pediatric ICU showed a slight decrease in adherence after the pandemic, from 67.51% (239/354) to 62.10% (262/422) (p = 0.115).

Considering professional categories, compliance remained stable among physicians (70.8% vs. 70.2%; p = 0.895). There was a non-significant increase among nurses (62.78% vs. 66.6%; p = 0.223) and nursing technicians (54.18% vs. 56.42%; p = 0.578). Among medical and nursing students, a non-significant reduction in HH compliance was identified (67.12% vs. 63.3%; p = 0.614). On the other hand, other healthcare professionals (including physiotherapists, pharmacists, and laboratory and radiology technicians) showed a statistically significant increase in HH compliance, from 61.78% to 75% (p = 0.012).

Before the pandemic, the most frequently used products for HH were non-medicated liquid soap and chlorhexidine (87%), with only 13% of healthcare workers opting for alcohol-based hand rubs. After the pandemic, a major shift was observed, with alcohol-based preparations accounting for 39% of HH events. This change was statistically significant (OR 4.30; 95% CI 3.32–5.58; p < 0.001). These findings are summarized in Table I.

**Table I.**
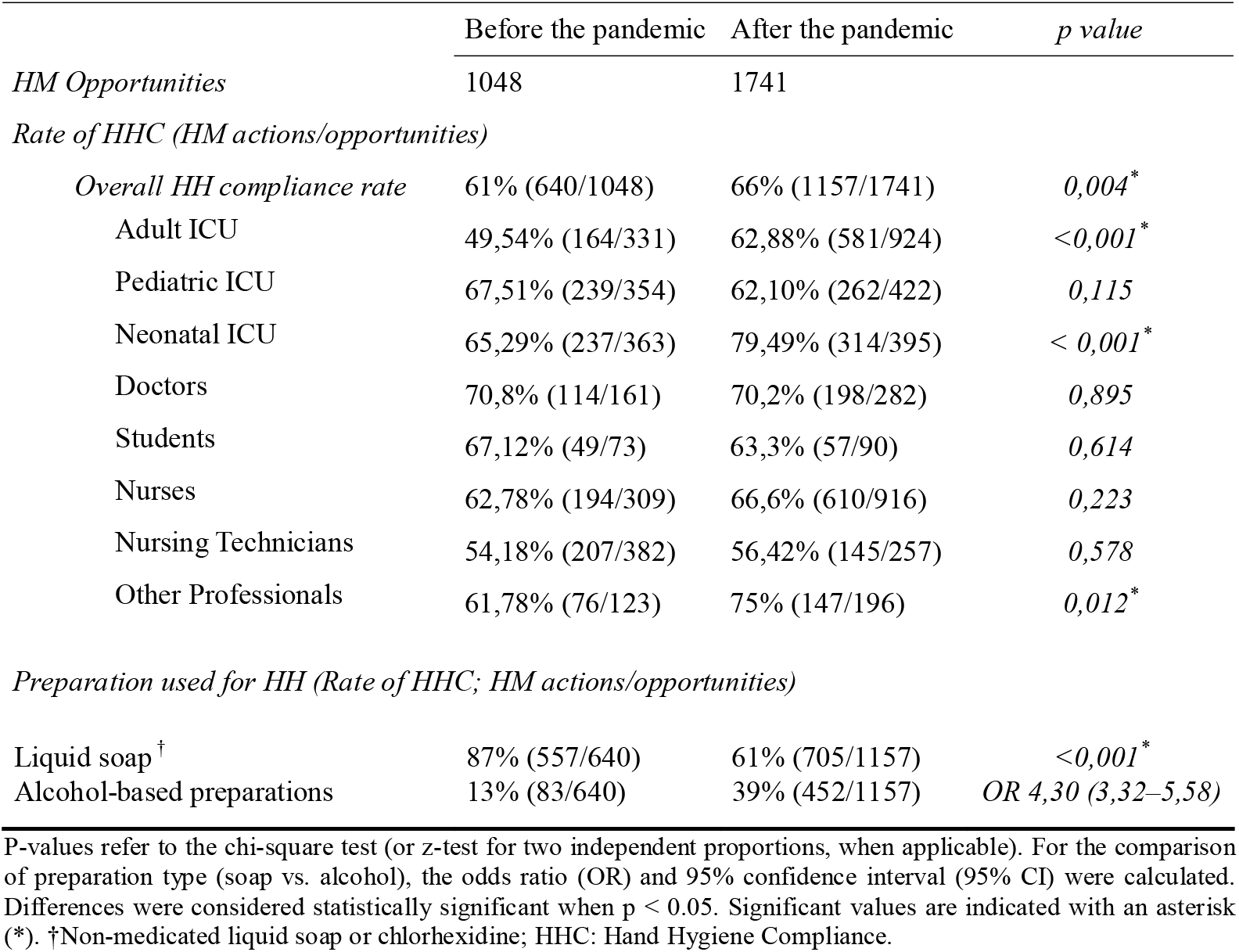
Comparison of Hand Hygiene Compliance Rate before and after the coronavirus 2019 (COVID-19) pandemic, according to professional category and preparation used.

|  | Before the pandemic | After the pandemic | *p value* |
| --- | --- | --- | --- |
| *HM Opportunities* | 1048 | 1741 |  |
| *Rate of HHC (HM actions/opportunities)* |  |  |  |
| *Overall HH compliance rate* | 61% (640/1048) | 66% (1157/1741) | 0,004* |
| Adult ICU | 49,54% (164/331) | 62,88% (581/924) | <0,001* |
| Pediatric ICU | 67,51% (239/354) | 62,10% (262/422) | 0,115 |
| Neonatal ICU | 65,29% (237/363) | 79,49% (314/395) | < 0,001* |
| Doctors | 70,8% (114/161) | 70,2% (198/282) | 0,895 |
| Students | 67,12% (49/73) | 63,3% (57/90) | 0,614 |
| Nurses | 62,78% (194/309) | 66,6% (610/916) | 0,223 |
| Nursing Technicians | 54,18% (207/382) | 56,42% (145/257) | 0,578 |
| Other Professionals | 61,78% (76/123) | 75% (147/196) | 0,012* |
| Liquid soap <sup>†</sup> | 87% (557/640) | 61% (705/1157) | <0,001* |
| Alcohol-based preparations | 13% (83/640) | 39% (452/1157) | OR 4,30 (3,32–5,58) |

P-values refer to the chi-square test (or z-test for two independent proportions, when applicable). For the comparison of preparation type (soap vs. alcohol), the odds ratio (OR) and 95% confidence interval (95% CI) were calculated. Differences were considered statistically significant when $p < 0.05$ . Significant values are indicated with an asterisk (\*). <sup>†</sup>Non-medicated liquid soap or chlorhexidine; HHC: Hand Hygiene Compliance.

Table II summarizes HH compliance after the pandemic according to the WHO’s “Five Moments” [6]. Marked variations were observed across ICUs. The compliance was significantly higher in the Neonatal ICU in Moments 1 (81.51%) and 2 (81.58%) compared with the Adult ICU (50.38% and 50.98%) and the Pediatric ICU (53.40% and 43.86%), with statistically significant differences (p < 0.001 for both). Pairwise comparisons (Fisher’s exact test with Holm adjustment) confirmed that, in both moments, the compliance was significantly higher in the Neonatal ICU compared with Adult and Pediatric ICUs. No significant differences were found between the Adult and Pediatric ICUs.

**Table II.** Hand Hygiene Compliance among Adult, Pediatric and Neonatal Intensive Care Units (ICUs) after the COVID-19 pandemic according to the WHO “5 Moments”.

| *Rate of HHC (%) and HM actions/opportunities* |  |  |  |  |
| --- | --- | --- | --- | --- |
|  | Adult ICU | Pediatric ICU | Neonatal ICU | *p* value |
| Moment 1 | 50,38% (132/262) | 53,40% (55/103) | 81,51% (97/119) | <0,001* |
| Moment 2 | 50,98% (52/102) | 43,86% (25/57) | 81,58% (31/38) | <0,001* |
| Moment 3 | 64,79% (92/142) | 64,62% (42/65) | 80,85% (38/47) | 0,1027 |
| Moment 4 | 80,80% (202/250) | 77,98% (85/109) | 86,55% (103/119) | 0,2230 |
| Moment 5 | 61,31% (103/168) | 62,50% (55/88) | 62,50% (45/72) | 0,9757 |
| *Preparation used for HH (Rate of HHC; HM actions/opportunities)* |  |  |  |  |
| Liquid soap <sup>†</sup> | 58,35% (339/581) | 63,74% (167/262) | 63,38% (199/314) | 0,193 |
| Alcohol-based | 41,65% (242/581) | 36,26% (95/262) | 36,62% (115/314) |  |

The World Health Organization (WHO) “Five Moments for Hand Hygiene”: (1) before touching the patient; (2) before performing a clean/aseptic procedure; (3) after risk of exposure to bodily fluids; (4) after touching the patient; (5) after touching surfaces near the patient. P-values refer to Pearson’s chi-square test (independence). Significant values are indicated with an asterisk (\*). The effect size (Cramer’s V) indicates a moderate effect at Moments 1 and 2 and a very low effect for soap vs. alcohol; <sup>†</sup>Non-medicated liquid soap or chlorhexidine; HHC: Hand Hygiene Compliance.

In Moment 3, the compliance across the three ICUs ranged from 64.62% to 80.85%, with no significant differences (p = 0.1027). In Moment 4, the compliance was globally high (77.98% to 86.55%), but without statistical difference (p = 0.2230). Finally, in Moment 5, the compliance was similar across ICUs, around 61% to 62.5%, with no significant difference (p = 0.9757). Regarding the type of product used for HH after the pandemic, no significant difference was found between ICUs in the proportion of liquid soap use (58.35%–63.74%) *versus* alcohol-based preparations (36.26%–41.65%) (p = 0.193).

## DISCUSSION

In recent decades, there has been a substantial increase in publications related to hand hygiene (HH), with growing evidence of improved compliance rates and reductions in healthcare-associated infections (HAIs) [14, 15]. During the COVID-19 pandemic, hand hygiene compliance (HHC) among healthcare professionals showed a significant increase compared with the pre-pandemic period, as a result of strengthened intervention measures aimed at improving compliance, including intensified educational efforts, greater access to alcohol-based formulations, and heightened awareness among healthcare workers regarding the risk of cross-transmission [16, 17, 18, 19]. However, some studies did not observe an increase in HHC after the pandemic, with potential contributing factors such as increased workload and the use of medical gloves instead of proper hand desinfection [20]. In our study, comparative analysis of HHC before and after the pandemic demonstrated a significant increase in overall compliance rate (p = 0.004). However, when discriminating among intensive care units (ICUs), we identified a significant increase in HHC in the Adult and Neonatal ICUs (p < 0.001), but this was not observed in the Pediatric ICU (p = 0.115). This result may be related to the perceived vulnerability of newborns in the Neonatal ICU and to preliminary evidence suggesting that children were less susceptible to *SARS-CoV-2* infection compared to adults during much of the pandemic period [21].

Considering professional categories, the compliance remained stable among physicians (p = 0.895), with a non-significant increase among nurses (p = 0.223) and nursing technicians (p = 0.578), and a non-significant decrease among medical and nursing students (p = 0.614). It is noteworthy that the reduction in the compliance among students may reflect limitations in the impact of interventions targeted at this group. Conversely, other healthcare professionals (physiotherapists, pharmacists, and laboratory and radiology technicians) showed a statistically significant increase in HHC (p = 0.012). In the multicenter PROHIBIT study [22], multilevel regression analysis among healthcare workers demonstrated that individual improvement was significantly associated with the nursing profession, lower activity index, and higher nurse-to-patient ratio. Indeed, an unfavorable nurse-to-patient ratio in our hospital may have contributed to lower HHC compared to physicians.

Among the various WHO initiatives, the “My Five Moments for Hand Hygiene” recommendation [6] encouraged healthcare professionals to increase HHC, improving compliance rates worldwide [23, 24, 25]. Based on the conceptual model of cross-transmission of microorganisms, it was designed for teaching and auditing purposes, emphasizing the use of alcohol-based formulations as the preferred method for HH [13]. In our study, the most frequently used products for HH before the pandemic were non-medicated liquid soap and chlorhexidine, and only 13% of healthcare professionals opted for alcohol-based preparations. After the pandemic, a significant shift occurred, with alcohol-based formulations chosen in 39% of HH events (p < 0.001). The “Odds Ratio” for alcohol-based preparation use after the pandemic was 4.30 (95% CI: 3.32–5.58), indicating more than a fourfold increase in the likelihood of alcohol use compared with the pre-pandemic period. This shift reflects compliance with WHO recommendations.

Analyzing the different scenarios regarding the WHO’s “Five Moments for Hand Hygiene” in the post-pandemic period, we identified significant differences among ICUs only in Moments 1 (before patient contact) and 2 (before aseptic procedures). The compliance was significantly higher in the Neonatal ICU at these moments compared with the Adult and Pediatric ICUs (p < 0.001 for both), once again reflecting the team’s heightened awareness of newborn vulnerability and the risk of severe infections in this population. Between the Adult and Pediatric ICUs, the compliance rates were similar, suggesting comparable practice patterns. In Moments 3, 4, and 5 (after exposure to body fluids, after patient contact, and after contact with patient surroundings), no significant differences were observed among ICUs (p > 0.10), with high compliance across all units. Notably, the compliance after patient contact (Moment 4) highlighted greater risk perception once exposure had already occurred. Other studies have reported similar findings [17, 18, 19, 26], suggesting that immediate risk, such as after exposure to body fluids or patient contact, favors greater compliance with HH practices across different settings. Regarding the type of preparation used, no significant differences were observed among ICUs after the pandemic (p > 0.19).

## CONCLUSION

The results reinforce that the COVID-19 pandemic led to an increase in healthcare workers’ hand hygiene compliance, although disparities persist among professional categories and hospital sectors. The use of alcohol-based preparations after the pandemic showed a significant increase, with more than a fourfold higher likelihood of alcohol use compared with the pre-pandemic period, in accordance with WHO recommendations. Overall, the findings indicate that HHC is more critical before patient contact (Moment 1) or before aseptic procedures (Moment 2), particularly in Adult and Pediatric ICUs; whereas the Neonatal ICU demonstrates superior performance across all moments, with significantly higher compliance in the two most critical moments. It can be concluded that there is a need for targeted educational strategies, with emphasis on adult and pediatric patient safety, and for the maintenance of institutional policies that encourage the use of alcohol-based preparations in daily practice.

## Data Availability

There are no legal or ethical restrictions.

## CONFLICT OF INTEREST AND FINANCING

This study has no conflict of interest and had no funding sources.

## Notes

### Competing Interest Statement

The authors have declared no competing interest.

### Funding Statement

Our work does not have funding.

### Author Declarations

The study is part of the project Strategies for reducing hospital costs and optimising the use of antimicrobials: rapid detection of resistance genes, synergism, and pharmacokinetic/pharmacodynamic analysis, phase 2 and was approved by the Permanent Research Ethics Committee Involving Human Beings of the State University of Maringa (CAAE 63610816.0.0000.0104).

## REFERENCES

1. Lai THT, Tang EWH, Fung KSC, Li KKW. Reply to “Does hand hygiene reduce SARS-CoV-2 transmission?”. Graefes Arch Clin Exp Ophthalmol. 2020;258(5):1135. doi: 10.1007/s00417-020-04653-4

2. Saitoh A, Sato K, Magara Y, Osaki K, Narita K, Shioiri K, et al. Improving hand hygiene adherence in healthcare workers before patient contact: a multimodal intervention in four tertiary care hospitals in Japan. J Hosp Med. 2020;15(5):262–7. doi: 10.12788/jhm.3446

3. Wang J, Feng H, Zhang S, Ni Z, Ni L, Chen Y, et al. SARS-CoV-2 RNA detection of hospital isolation wards hygiene monitoring during the coronavirus disease 2019 outbreak in a Chinese hospital. Int J Infect Dis. 2020;94:103–6. doi: 10.1016/j.ijid.2020.04.024

4. Centers for Disease Control and Prevention (CDC). Comprehensive hospital preparedness checklist for coronavirus disease 2019 (COVID-19). Atlanta: US Department of Health and Human Services; 2020. Accessed on: https://www.cdc.gov/coronavirus/2019-ncov/downloads/HCW_Checklist_508.pdf

5. Boyce JM, Pittet D; Healthcare Infection Control Practices Advisory Committee; HICPAC/SHEA/APIC/IDSA Hand Hygiene Task Force. Guideline for hand hygiene in health-care settings. MMWR Recomm Rep. 2002;51(RR-16):1–45, quiz CE1-4. doi: 10.1086/503164

6. World Health Organization. WHO guidelines on hand hygiene in health care: first global patient safety challenge: clean care is safer care. Geneva: WHO; 2009. Accessed on: https://www.who.int/publications/i/item/9789241597906

7. European Centre for Disease Prevention and Control. Point prevalence survey of healthcare-associated infections and antimicrobial use in European acute care hospitals. Stockholm: ECDC; 2013. Accessed on: https://www.ecdc.europa.eu/en/publications-data/point-prevalence-survey-healthcare-associated-infections-and-antimicrobial-use

8. Magill SS, Edwards JR, Bamberg W, Beldavs ZG, Dumyati G, Kainer MA, et al. Multistate point-prevalence survey of health care-associated infections. N Engl J Med. 2014;370(13):1198–208. doi: 10.1056/NEJMoa1306801

9. Brasil. Agência Nacional de Vigilância Sanitária (ANVISA). Segurança do paciente: higienização das mãos. Brasília: Ministério da Saúde; 2009. Accessed on: https://bvsms.saude.gov.br/bvs/publicacoes/seguranca_paciente_servicos_saude_higienizacao_maos.pdf

10. World Health Organization. WHO guidelines on hand hygiene in health care (advanced draft): a summary. Clean hands are safer hands. Geneva: WHO; 2005. Accessed on: https://apps.who.int/iris/handle/10665/69579

11. Pittet D, Allegranzi B, Storr J. The WHO Clean Care is Safer Care programme: fieldtesting to enhance sustainability and spread of hand hygiene improvements. J Infect Public Health. 2008;1:4–10. doi: 10.1016/j.jiph.2008.08.002

12. World Health Organization. Tools for institutional safety climate. Geneva: WHO; 2009. Accessed on: https://www.who.int/gpsc/5may/tools/safety_climate/en/

13. Loftus MJ, Guitart C, Tartari E, Stewardson AJ, Amer F, Bellissimo-Rodrigues F, et al. Hand hygiene in low- and middle-income countries. Int J Infect Dis. 2019;86:25–30. doi: 10.1016/j.ijid.2019.06.002

14. Allegranzi B, Pittet D. Role of hand hygiene in healthcare-associated infection prevention. J Hosp Infect. 2009;73:305–15. doi: 10.1016/j.jhin.2009.04.019

15. Kingston L, O’Connell NH, Dunne CP. Hand hygiene-related clinical trials reported since 2010: a systematic review. J Hosp Infect. 2016;92(4):309–20. doi: 10.1016/j.jhin.2015.11.012

16. Kong A, Botero Suarez CS, Rahamatalli B, Shankweiler J, Karasik O. Hand hygiene and hospital-acquired infections during COVID-19 increased vigilance: one hospital’s experience. HCA Healthc J Med. 2021;2(5):379–84. doi: 10.36518/2689-0216.1296

17. Wang Y, Yang J, Qiao F, Feng B, Hu F, Xi ZA, et al. Compared hand hygiene compliance among healthcare providers before and after the COVID-19 pandemic: a rapid review and meta-analysis. Am J Infect Control. 2022;50(5):563–71. doi: 10.1016/j.ajic.2021.11.030

18. Guerrero-Soler M, Gras-Valentí P, Gómez-Sotero IL, et al. Impact of COVID-19 on the degree of compliance with hand hygiene: a repeated cross-sectional study. Epidemiol Infect. 2024;152:e69. doi: 10.1017/S0950268824000505

19. Zhang X, Ma Y, Kong L, Li Y, Wang J, Li N, et al. The impact of COVID-19 pandemic on hand hygiene compliance of healthcare workers in a tertiary hospital in East China. Front Med (Lausanne). 2023;10:1160828. doi: 10.3389/fmed.2023.1160828

20. Sandbøl SG, Glassou EN, Ellermann-Eriksen S, Haagerup A. Hand hygiene compliance among healthcare workers before and during the COVID-19 pandemic. Am J Infect Control. 2022;50(7):719–23. doi: 10.1016/j.ajic.2022.03.014

21. Viner RM, Mytton OT, Bonell C, Melendez-Torres GJ, Ward J, Hudson L, et al. Susceptibility to SARS-CoV-2 infection among children and adolescents compared with adults: a systematic review and meta-analysis. JAMA Pediatr. 2021;175(2):143–56. doi: 10.1001/jamapediatrics.2020.4573

22. van der Kooi T, Sax H, Grundmann H, Pittet D, de Greeff S, van Dissel J, et al; PROHIBIT consortium. Hand hygiene improvement of individual healthcare workers: results of the multicentre PROHIBIT study. Antimicrob Resist Infect Control. 2022;11(1):123. doi:10.1186/s13756-022-01148-1. Erratum in: Antimicrob Resist Infect Control. 2023;12(1):13. doi: 10.1186/s13756-022-01148-1

23. Pittet D, Allegranzi B, Boyce J. The World Health Organization guidelines on hand hygiene in health care and their consensus recommendations. Infect Control Hosp Epidemiol. 2009;30(7):611–22. doi: 10.1086/600379

24. Allegranzi B, Gayet-Ageron A, Damani N, Bengaly L, McLaws ML, Moro ML, et al. Global implementation of WHO’s multimodal strategy for improvement of hand hygiene: a quasi-experimental study. Lancet Infect Dis. 2013;13(10):843–51. doi: 10.1016/S1473-3099(13)70163-4

25. Tschudin-Sutter S, Sepulcri D, Dangel M, Schuhmacher H, Widmer AF. Compliance with the World Health Organization hand hygiene technique: a prospective observational study. Infect Control Hosp Epidemiol. 2015;36(4):482–3. doi: 10.1017/ice.2014.82

26. Anwar MM, Elareed HR. Improvement of hand hygiene compliance among health care workers in intensive care units. J Prev Med Hyg. 2019;60(1):E31–E35. doi: 10.15167/2421-4248/jpmh2019.60.1.918

